# Ancestry-specific effects of *APOE* on Alzheimer Disease Endophenotypes

**DOI:** 10.64898/2026.03.23.26349095

**Authors:** Ana I. Boeriu, Tianjie Gu, Brian Fullton-Howard, Esteban M Lucero, Paras Garg, Andrew J. Sharp, Jonathan A. Shortt, Chris R. Gignoux, Farid Rajabli, Anthony J. Griswold, Kristine Yaffe, Shea J. Andrews

**Affiliations:** Department of Psychiatry and Behavioral Sciences, University of California, San Francisco, CA, USA; John P. Hussman Institute for Human Genomics, University of Miami, Miami, Florida, US; Department of Genetics and Genomic Sciences, Icahn School of Medicine at Mount Sinai, New York, NY, USA; Ronald M. Loeb Center for Alzheimer’s Disease, Icahn School of Medicine at Mount Sinai, New York, NY, US; Department of Biomedical Informatics, University of Colorado Anschutz Medical Campus, Aurora, CO, USA; Department of Genetics and Genomic Sciences and Mindich Child Health and Development Institute, Icahn School of Medicine at Mount Sinai, New York, NY, USA; Dr. John T Macdonald Foundation Department of Human Genetics, University of Miami, Miami, Florida, USA; Department of Neurology, and Department of Epidemiology, University of California, San Francisco, CA, USA

**Keywords:** APOE, Alzheimer’s disease, Genetic ancestry, Local ancestry, Plasma biomarkers, cognition, pTau217, Amyloid-β

## Abstract

**INTRODUCTION:** *APOE*\*ε4 is the strongest genetic risk factor for Alzheimer’s disease (AD), yet its effect varies across ancestral populations. As plasma biomarkers increasingly inform AD diagnosis, failure to account for both *APOE* genotype and genetic ancestry could lead to diagnostic misclassification.

**METHODS:** In 2,733 participants from the Health and Aging Brain Study–Health Disparities (HABS-HD), we evaluated whether genetically-inferred ancestry modifies *APOE* associations with AD endophenotypes. Regression models were applied across three frameworks: ancestry-stratified analyses, compositional modeling of global ancestry, and local ancestry analyses at the *APOE* locus.

**RESULTS:** *APOE* ε4+ was associated with higher pTau_217_ and pTau_181_, lower Aβ_42_/Aβ_40_, and worse global cognition (CDR-SB), significant within ancestry groups after FDR correction. The ε4+ effect on pTau_217_ was ∼2.5-fold larger in EUR-like than AFR-like participants.

**DISCUSSION:** African ancestry attenuates *APOE*\*ε4+ associations with pTau_217_, highlighting the need to consider both *APOE* genotype and ancestry in biomarker-based AD evaluations.

## 1. Background

Alzheimer’s disease (AD) affects over 7 million Americans, with projections reaching nearly 14 million by 2060^1^ – a burden that will fall disproportionately on underrepresented racial and ethnic groups. Black and Hispanic populations are approximately 1.5 and 2 times more likely to develop dementia compared with non-Hispanic White older adults^2,3^. Additionally, Black individuals show reduced cortical thickness when amyloid-positive^4^ and lower CSF concentrations of total tau and p-tau ^5^. These differences in population risk can be attributed to both genetic and environmental risk factors.

Among genetic contributors, the apolipoprotein E (*APOE*) gene is the strongest genetic risk factor for late-onset AD^6,7^, with ε2, ε3, and ε4 alleles exhibiting dose-dependent associations with AD risk. Relative to the most common ε3 allele, the ε4 allele is associated with increased risk of AD and an earlier age of onset, with one ε4 allele increasing risk 3-4-fold and two ε4 alleles 9-15-fold^8^; ε4 homozygotes show symptom onset at a mean age of 65 years, earlier than in e3 alleles^9^. In contrast, the ε2 allele confers protection and delays disease onset^10^.

Although *APOE* ε4 is the strongest genetic risk factor for late-onset AD, its estimated association with AD risk varies across populations^6^. This variation may reflect genetic architecture, environmental, or social factors that differ across populations or their interactions. Compared with race/ethnicity, genetic ancestry more directly captures differences in genetic architecture that may modify APOE effects, motivating evaluation of APOE associations across continental, global, and local ancestry^11,12^. Genetic ancestry, inferred from an individual’s genetic similarity to reference populations, can be characterized at varying resolutions^13–15^. At the broadest level, continental ancestry assigns each individual to a single group based on their predominant genetic similarity to a reference population^12^. Rather than a single assignment, global ancestry quantifies each individual’s ancestry across the genome as continuous proportions derived from multiple reference populations^13,14^. At the chromosomal level, local ancestry identifies the ancestral origin of specific genomic segments, assigning ancestry to each haplotype at a given locus and capturing ancestry-specific genetic variation across the genome^13,14^. These levels – continental group, global proportion, and local ancestry– describe ancestry at increasingly fine resolution, and associations may vary across these levels. We therefore examined all three to capture distinct aspects of *APOE*-related AD risk. At the population group level, the ε4 allele confers the highest AD risk in East Asian ancestry populations, followed by European, African, and Amerindian ancestry, while ε2 shows attenuated protective effects in African and Amerindian populations^6,16^. At the global level, higher Amerindian ancestry protects against ε4-related cognitive decline^17^, while higher African ancestry is associated with worse functional cognitive scores in *APOE* ε4 carriers with severe neuritic plaque burden^18^. At the *APOE* locus, African local ancestry reduces AD risk independent of *APOE* genotype, with ε4 alleles on African-derived haplotypes conferring lower risk than those on European haplotypes^13,15^.

While genetic ancestry moderates the effect of *APOE* on AD risk, few studies have systematically examined how genetic ancestry modifies *APOE* effects on AD endophenotypes beyond clinical or cognitive diagnosis^6,17^. Leveraging the Health and Aging Brain Study–Health Disparities (HABS-HD) cohort, we evaluated *APOE* effects on blood-based biomarkers, cognitive performance, and brain morphometry using (1) ancestry-stratified regression to assess *APOE* associations within continental ancestry groups, (2) *APOE*-by-global ancestry interaction models, and (3) local ancestry analysis at the *APOE* locus to examine *APOE* effects among homozygous local ancestry groups and test for *APOE*-by-local ancestry interactions.

## 2. Methods

### 2.1 Study Population

Clinical and genomic data were obtained from HABS-HD (data release 6), an ongoing community-based study examining brain health and Alzheimer’s disease in the Dallas-Fort Worth, Texas area^19^. Participants were adults aged ≥ 30 years who self-reported as non-Hispanic White, Mexican American, or African American; detailed eligibility criteria have been published previously^19^. All participants underwent neuropsychological testing, blood collection, and MRI^19–21^. The study protocol is approved by the North Texas Regional Institutional Review Board^19^.

Among 3,838 participants at baseline, we sequentially excluded 1,105 individuals for age < 55 years (n = 515), failed neuroimaging quality control (n = 16), missing genome-wide ancestry data (n = 567), assignment to EAS-like ancestry (n = 2) or Oceanic-like ancestry proportions greater than 5% (n = 1), or missing *APOE* genotypes (n = 4). The age exclusion criterion was based on prior studies demonstrating age-dependent *APOE* effects^6,22^. Plasma pTau measurements (data release 7) were merged with the analytic dataset using participant identifiers and did not alter cohort composition. The final sample consisted of 2,733 participants.

### 2.2 AD Endophenotypes

#### 2.2.1 Cognitive Function

Participants completed a neuropsychological battery^23,24^ including the Mini-Mental State Examination (MMSE), the Clinical Dementia Rating Scale (CDR), Trail Making Test Parts A and B (TMT-A & B), Wechsler Memory Scale–III (WMS-III) Logical Memory I and II, the Spanish English Verbal Learning Test (SEVLT)^25^, WMS-III Digit Span, Letter Fluency (FAS), and the Animal Naming Test (ANT). Demographically adjusted cognitive composite scores for memory, executive function, and verbal ability were calculated by stratifying participants by age (≤65, ≥66 years), education (0–7, 8–12, ≥13 years), and primary language (English, Spanish), converting raw scores to z-scores within each stratum, then averaging z-scores across tests within each domain: memory (WMS-III Logical Memory I and II, SEVLT), executive function (WMS-III Digit Span, TMT-A and TMT-B), and verbal ability (FAS, ANT)^21^ (Figure S1). Global cognitive performance was assessed using MMSE and CDR Sum of Boxes (CDR-SB) as separate outcomes.

#### 2.2.2 Blood-based Biomarkers of AD pathology

Plasma biomarkers for amyloid (Aβ_42_/Aβ_40_), tau (pTau_181_, pTau_217_, total tau), and neurodegeneration (NfL) were assayed using Simoa on the HD-X platform (Quanterix, Billerica, MA), and coefficients of variation were ≤ 5% for all assays^26^. Assays were run across two kit versions (r3 and r5), requiring a harmonization^27^ pipeline to correct systematic batch effects. Raw concentrations were log₁₀-transformed, outliers were removed using the interquartile range method (Q1 − 1.5×IQR; Q3 + 1.5×IQR), and log₁₀ values were z-score standardized within each kit batch. For participants with multiple analytical runs, the z-score from the earliest run was retained; participants missing all biomarker measurements were excluded (Figure S2). Plasma pTau_217_ was measured using the Quanterix PLUS lyophilized assay (data release 7), released as a single analytical batch; values were log₁₀-transformed and z-score standardized without batch harmonization.

#### 2.2.3 Neuroimaging

Structural T1-weighted MRI scans were processed as previously described^19^. Cortical thickness was computed as a single composite, the surface-area-weighted average across an AD-signature meta-region of interest (entorhinal cortex, fusiform, inferior temporal, and middle temporal gyri)^28^ and standardized to z-scores. Hippocampal volumes and intracranial volume (ICV) were derived using HippoDeep^29^; bilateral hippocampal volumes were averaged and standardized to z-scores.

### 2.3 Genetics

#### 2.3.1 APOE genotyping

*APOE*-defining SNV rs429358 and rs7412 were directly genotyped using TaqMan assays^30^. *APOE* genotypes were categorized into three risk groups: ε2+ carriers (ε2/ε3, ε2/ε2), ε3/ε3 (reference), and ε4+ carriers (ε2/ε4, ε3/ε4, ε4/ε4), with ε2/ε4 categorized as ε4+ due to increasing AD risk^10,31^. Individuals heterozygous at both SNVs were assigned ε2/ε4, as the alternative ε1/ε3 diplotype requires the ε1 haplotype, which is extremely rare^32^.

#### 2.3.2 Genome-wide genotyping and imputation

DNA extracted from peripheral blood buffy coat samples was genotyped using the Illumina Global Screening Array (GSA) and underwent quality control for call-rate, Hardy-Weinberg equilibrium, and cryptic relatedness. Imputation was then performed on the NIH Imputation Server with the TOPMed r3 panel^33^, yielding ∼22 million high-quality SNPs across 3,232 samples (Supplementary Methods).

#### 2.3.3 Reference Panel

All genetic similarity analyses used 1000 Genomes Project (1KG) ^34^ and Human Genome Diversity Project (HGDP)^35^ reference samples from gnomAD v3.1.2, spanning five superpopulations: African (AFR, n = 637), Admixed American (AMR, n = 337), East Asian (EAS, n = 498), European (EUR, n= 510), and South Asian (SAS, n=481) (Figures S3-S5).

#### 2.3.4 Genetic Inferred Population Groups

Participant genotypes were projected onto the principal component space derived from the reference panel, and each individual was classified to the nearest of five superpopulations based on the Euclidean distance across the first 10 PCs^35,36^. Participants assigned to the EAS-like group were excluded due to small sample sizes (n=2); no participant was classified as SAS-like. Analyses resulted in three groups: AFR-like, AMR-like, and EUR-like. These labels denote genetic similarity to reference populations rather than lines of descent (Supplementary Methods).

#### 2.3.5 Global Ancestry Proportions

Ancestry proportions were estimated using ADMIXTURE in unsupervised mode (K=12, selected at the plateau in cross-validation error, beyond which additional components provided minimal improvement [Figure S6]) on LD-pruned variants shared between HABS-HD and the reference panel (Figure 1). Each cluster was labeled by the reference superpopulation with the highest mean proportion and estimates from clusters sharing the same label were then pooled to obtain continuous ancestry proportions per participant. Residual EAS-like proportions were pooled into AMR-like and residual SAS-like proportions into EUR-like for downstream analyses, as East Asian and South Asian contributions were minimal and reflect shared deep ancestral components rather than recent population-specific gene flow^35,37^ (Supplementary Methods).

**Figure 1:**
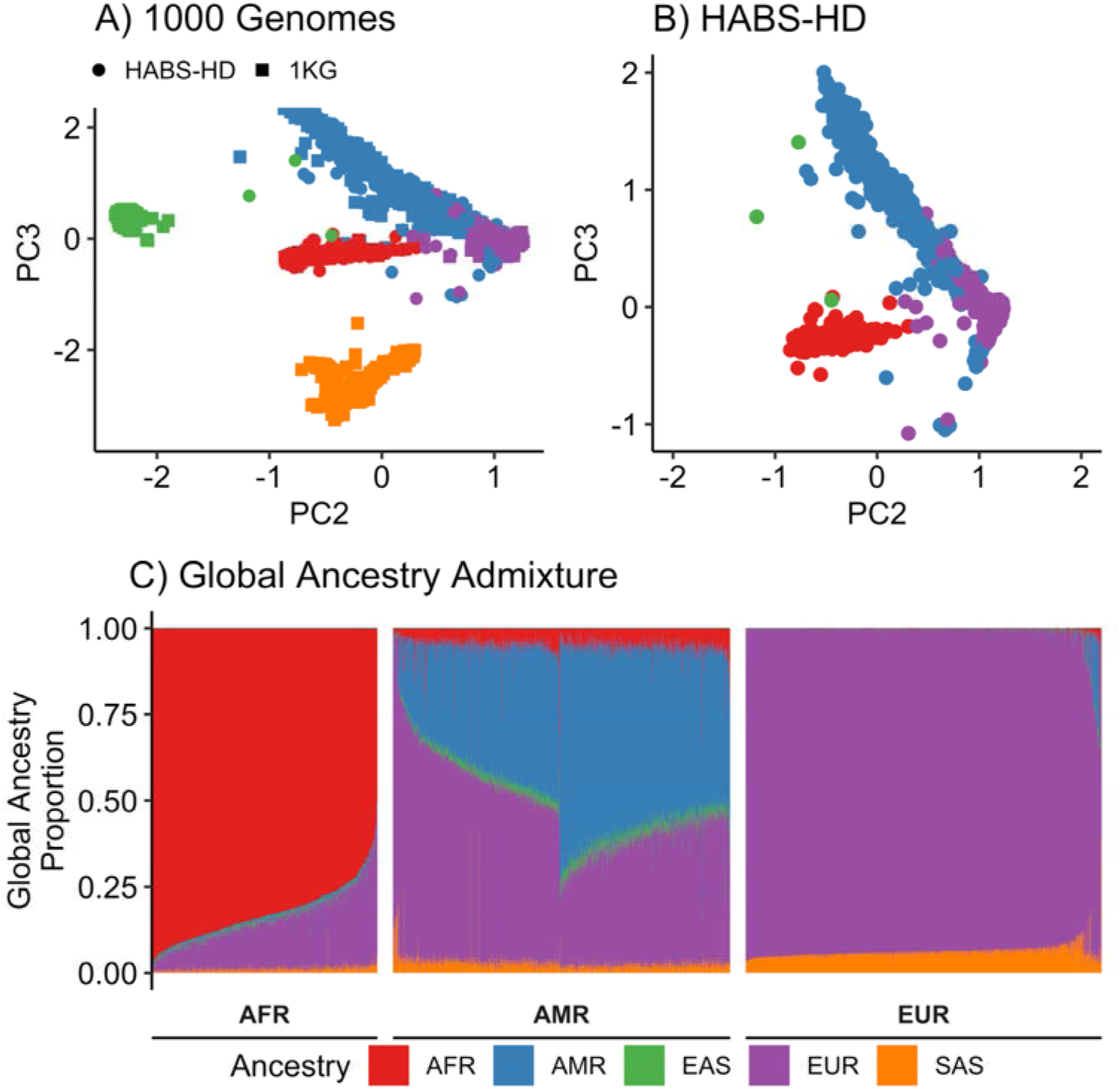
Genetic Ancestry Characterization of Study Participants. (A) Principal component analysis (PCA) of HABS-HD samples with 1000 Genomes Project reference populations demonstrating continental ancestry clustering. (B) PCA of HABS-HD cohort (N=2,733) showing genetic structure and ancestry group distribution: EUR (purple, n=1,138), AMR (blue, n=953), and AFR (red, n=642). (C) Global ancestry admixture proportions by continental ancestry group, displaying varying degrees of ancestral contributions within each group. AFR, African ancestry; AMR, Admixed American ancestry; EAS, East Asian ancestry; EUR, European ancestry; PC, principal component; SAS, South Asian ancestry.

#### 2.3.6 Local Ancestry Inference at the APOE Locus

Haplotype-specific genetic similarity was inferred using G-Nomix^38^ on phased genotype data within a 1Mb region centered on the *APOE* locus (chr19: 44,405,791 – 45,409,393 hg38) using a reference panel of five superpopulations (AFR-like, AMR-like, EUR-like, SAS-like, EAS-like; Figure S7). Given the minimal East Asian and South Asian ancestry in HABS-HD (35 individuals with at least one EAS or SAS assigned haplotype, 1.3% of the cohort), EAS-like was collapsed into AMR-like, and SAS-like into EUR-like, as they are not reliably distinguished at a single locus^35^, yielding three genetically similar groups (AFR-like, AMR-like, and EUR-like). This relabeling affects 21 of the 1,328 participants in the analytical strata (Table S1). Analyses were then restricted to participants with concordant local ancestry across both APOE haplotypes (AFR-like/AFR-like, EUR-like/EUR-like, or AMR-like/AMR-like) to ensure that *APOE* effects could be attributed to a single ancestral background; individuals with discordant segment-level assignments were classified as mixed local ancestry and excluded. To isolate ε4-specific effects, ε2-carrier genotypes were excluded, and the remaining genotypes were collapsed into ε3/ε3 (reference), ε3/ε4, ε4/ε4 (ε4+) (Table S1; Supplemental Material)

### 2.4 Statistical Analysis

Baseline characteristics were compared across genetic similarity groups using Kruskal-Wallis tests for continuous variables and Pearson’s chi-square tests for categorical variables (Table 1). Pairwise comparisons used Bonferroni-corrected t-tests and chi-square tests (Table S2). To assess how genetic ancestry modifies *APOE* effects, we evaluated *APOE* associations with cognitive function (MMSE, CDR-SB, memory, executive function, verbal ability), plasma biomarkers (Aβ_42/40_, pTau_181_, pTau_217_, total tau, NfL), and brain morphometry (cortical thickness, hippocampal volume) using three complementary approaches. First, *APOE* ε2+ and ε4+ effects were estimated within each genetic similarity group and the full cohort using ancestry-stratified linear regression, with pairwise Wald tests comparing coefficients between groups^39,40^. Second, *APOE*-by-global ancestry interactions were tested using compositional linear regression with isometric log-ratio (ILR) transformation to account for the sum-to-one constraint of ancestry proportions. Because ancestry proportions are compositional — non-negative and summing to one for each participant — they cannot all be entered directly into standard regression, as the sum to one constraint includes perfect collinearity with the intercept; the ILR transformation maps them into two unconstrained coordinates suitable for linear modeling^41^. Under the primary parameterization (EUR-like, AFR-like, AMR-like), the two ILR coordinates are defined as EUR-like versus combined AFR-like and AMR-like (ILR1) and AFR-like versus AMR-like (ILR2). Alternative parameterizations were also evaluated as sensitivity analyses (Supplementary Methods). Third, *APOE* ε4+ effects by local ancestry at the *APOE* locus were examined using two complementary approaches: ancestry-stratified models within homozygous groups (AFR/AFR, EUR/EUR, AMR/AMR) and interaction models in the full cohort. Local ancestry models used the first three principal components in place of global ancestry proportions to avoid overadjustment for ancestry.

Benjamini-Hochberg false-discovery-rate correction was applied within each pre-specified family of tests: continental within-group APOE effects (96 tests), continental pairwise contrasts between groups (72 tests), global-ancestry APOE × ILR interactions (primary parameterization, 48 tests), local-ancestry within-stratum APOE effects (48 tests), and local-ancestry APOE × ancestry interactions (24 tests). Alternative ILR parameterizations were treated as sensitivity analyses, and meta-analysis replication of pre-specified effects was treated as confirmatory; neither was FDR-corrected. Throughout, we use “attenuated” only when a reduction in the estimated effect is supported by a formal test at p < 0.05, not when point estimates simply differ.

To assess generalizability, inverse-variance-weighted random-effects meta-analyses (restricted maximum likelihood [REML]) combined HABS-HD with an independent multi-ancestry cohort^42^ (n = 2086; AFR-like, EUR-like participants) for Aβ_42/40_, pTau_181_, and total tau across the ancestry-stratified and global-ancestry-proportion frameworks. Given only two cohorts, we focused on the consistency of effect direction and the significance of the pooled estimates rather than heterogeneity statistics. An effect was considered replicated only if the estimate in the independent cohort was directionally consistent with HABS-HD and independently significant; the significance of the pooled estimate alone was not treated as replication.

All models included age, sex, and genetic ancestry as covariates, with *APOE* genotype as the primary predictor. Domain-specific covariates were BMI and eGFRcr (2021 CKD-EPI equation without race^43^), for plasma biomarkers; MRI scanner and intracranial volume for neuroimaging; and education and primary language for cognitive assessments. Covariates were selected a priori to account for established phenotype-specific confounding and measurement effects. Cardiometabolic comorbidities (hypertension, dyslipidemia, and diabetes) were not adjusted for as they potentially lie on the causal pathway between *APOE*- or ancestry-associated pathways and could result in over-adjustment. Missing BMI (0.7%) and creatinine (2%) values were imputed using random forest imputation, while participants with missing outcome data were excluded. All analyses were performed in R version 4.5.2 with α = 0.05 (Supplementary Methods).

## 3. Results

### 3.1 Baseline Characteristics

The final cohort comprised 2,733 participants (mean age 67 years; 61% female) spanning the cognitive spectrum across three genetically inferred population groups: 1,138 EUR-like (41.64%), 642 AFR-like (23.48%), and 953 AMR-like (34.86%) participants (Table 1, Figure 1). *APOE* ε4+ carrier frequency was highest in AFR-like (42%), followed by EUR-like (30%) and AMR-like (18%) participants. EUR-like participants were older with higher cognitive performance across all domains, while both AMR-like participants and AFR-like participants had lower pTau_181_ and pTau_217_ levels than EUR-like participants (Table S2). Genetically inferred ancestry corresponded closely with self-reported ethnicity (2,668/2,733, 97.6%; Table 1): 98.5% of self-reported White participants were assigned EUR-like, 99.5% of Black participants AFR-like, and 95.4% of Hispanic participants AMR-like. Discordant assignments were mostly self-reported Hispanic participants with predominantly European (n=38) or African (n=7) genetic ancestry.

### 3.2 *APOE* ε4 Effects on AD Endophenotypes Vary by Genetically Inferred Population Groups

In the entire cohort, ε4+ was associated with lower Aβ_42_/Aβ_40_ (β=−0.42, 95% CI −0.51 to −0.33), higher pTau_217_ (β=0.55, 95% CI 0.46 to 0.64), higher pTau_181_ (β=0.31, 95% CI 0.23 to 0.40), worse performance across all cognitive domains, and reduced hippocampal volume and cortical thickness. ε2+ was associated with lower pTau_217_ (β=−0.17, 95% CI −0.29 to −0.05) but showed limited associations with other endophenotypes (Figure 2, Table S3).

**Figure 2:**
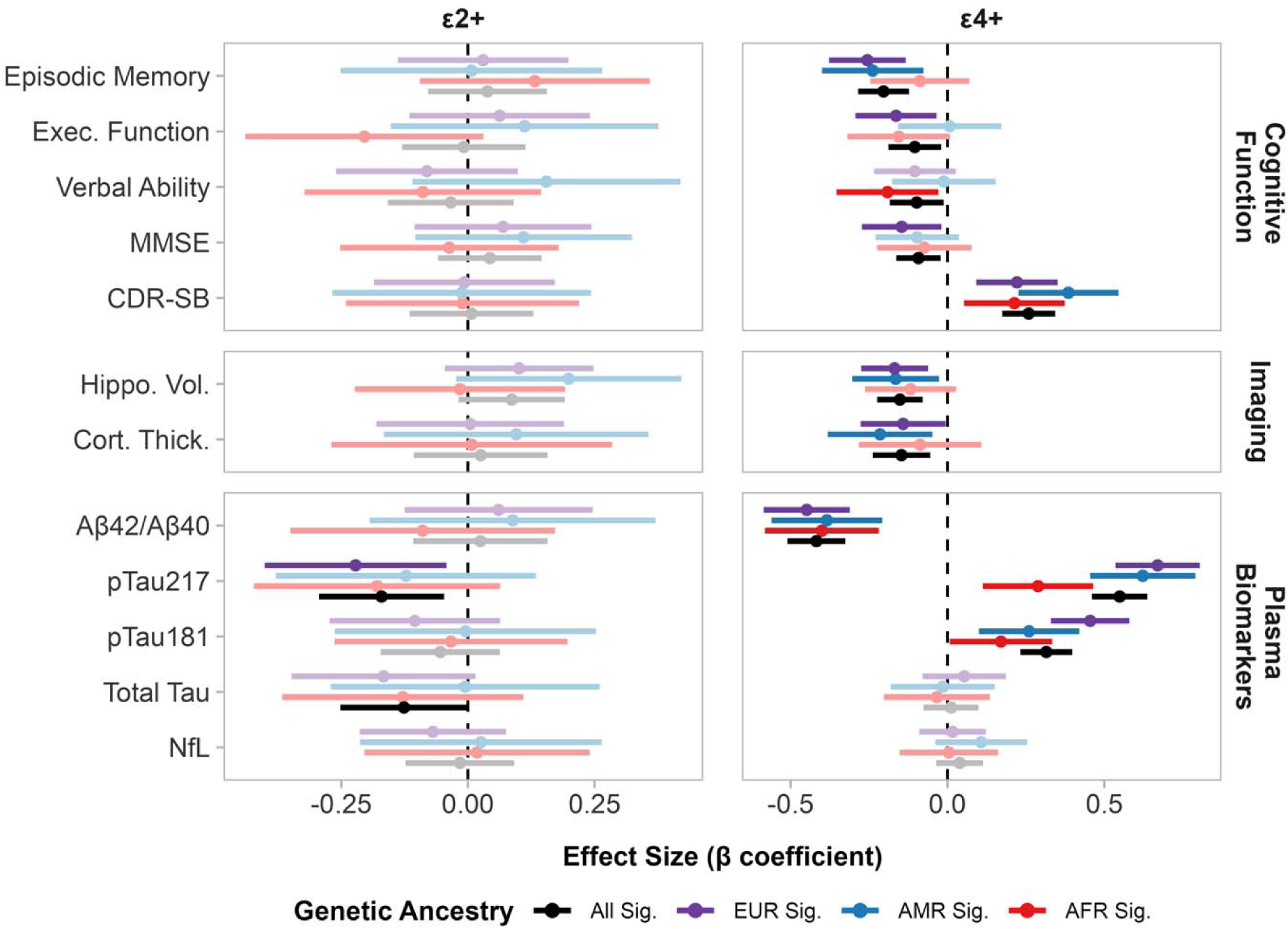
Association of *APOE* with AD endophenotypes by genetically inferred population groups. Forest plot showing standardized effect sizes (β coefficients) with 95% confidence intervals for *APOE* ε4+ and ε2+ risk groups compared to ε3/ε3 reference group. Results are stratified by genetically inferred population groups, and colored points indicate statistically significant associations (95% CI does not cross 0 and p value < 0.05). Standardized effects allow direct comparison of effect magnitudes across different outcome domains. Color indicates nominal statistical significance (p-value < 0.05, uncorrected). FDR-adjusted q-values for all tests are reported in Table S3.

In ancestry-stratified analyses, ε4+ effects on pTau_217_ were approximately 2.5-fold larger in EUR-like (β=0.68, 95% CI 0.55 to 0.82) than AFR-like participants (β=0.28, 95% CI 0.11 to 0.44), a difference that survived FDR correction (Table S4). AMR-like participants showed intermediate effects (β=0.60, 95% CI 0.44 to 0.76). A directionally similar but FDR non-significant pattern was observed for pTau_181_: EUR-like (β=0.44, 95% CI 0.32 to 0.56) versus AFR-like (β=0.17, 95% CI 0.01 to 0.34). In contrast, ε4+ effects on Aβ_42_/Aβ_40_ were consistent across groups (EUR-like β=−0.45; AFR-like β=−0.42; AMR-like β=−0.37). The ε2+ protective effect on pTau_217_ was similarly driven by EUR-like participants.

For cognition, ε4+ was associated with worse CDR-SB in all three groups, worse MMSE, episodic memory, and executive function in EUR-like participants, worse episodic memory in AMR-like participants, and worse verbal ability in AFR-like participants. For brain morphometry, ε4+ was associated with reduced cortical thickness and hippocampal volume in EUR-like and AMR-like but not AFR-like participants. Between-group differences were not significant for cognition or neuroimaging, and ε2+ showed no significant associations with either domain (Table S3, Table S4). After FDR correction (96 tests; Table S3), ε4+ effects on pTau217, Aβ_42_/Aβ_40_ and CDR-SB remained significant in all three ancestry groups. pTau181 and episodic memory survived in EUR-like and AMR-like participants, hippocampal volume in EUR-like, and cortical thickness in AMR-like; no imaging association survived in AFR-like participants. The ε2+ effect on pTau217 survived in the full cohort only.

In meta-analysis with an independent multi-ancestry cohort, the ε4+ effect on pTau_181_ in EUR-like participants replicated – directionally consistent and independently significant in both cohorts – and remained significant when pooled (pooled β=0.42, 95% CI 0.31 to 0.52). In AFR-like participants, the effect was directionally consistent across cohorts and significant when pooled (pooled β=0.14, 95% CI 0.01 to 0.27), but did not meet our replication criterion as it was not independently significant in the replication cohort (β=0.08, 95%CI -0.14 to 0.30). The pooled analysis did not test whether the EUR-like and AFR-like effects differ (Figure S8, Table S5).

### 3.3 Global Ancestry Proportions Moderate *APOE* ε4+ Effects on AD Endophenotypes

Using the primary ILR parameterization, ILR1 contrasts European ancestry with the combined African and AMR ancestry proportions, while ILR2 contrasts African and AMR ancestry. Under this parameterization, the sign of the interaction coefficient indicates the direction of the *APOE* × ancestry association: positive ILR1 coefficients indicate stronger *APOE* associations with increasing European ancestry, whereas positive ILR2 coefficients indicate stronger *APOE* associations with increasing African relative to AMR ancestry (and negative coefficients indicate the converse). ε4+ effects on pTau_181_ (β=0.045, 95% CI 0.013 to 0.077) and pTau_217_ (β=0.038, 95% CI 0.003 to 0.072) strengthened with increasing EUR proportion relative to combined AFR and AMR proportions. Thus, a positive ILR1 interaction coefficient indicates stronger ε4+ associations with increasing EUR-like ancestry relative to combined AFR and AMR proportions. In the AFR versus AMR contrast, ε4+ effects on pTau_217_ were attenuated with increasing AFR proportion relative to AMR (β=−0.058, 95% CI −0.099 to −0.017). No significant ε2+ or ε4+ × ancestry interactions were observed for Aβ_42_/Aβ_40_, total tau, or NfL. For cognition, ε4+ effects on verbal ability differed between AFR and AMR proportions (β=−0.037, 95% CI −0.071 to −0.002), and ε2+ protective effects on executive function were attenuated with increasing EUR proportion (β=0.042, 95% CI 0.003 to 0.081). No significant interactions were observed for neuroimaging (Figure 3, Table S6). No APOE × ancestry interaction in this framework survived FDR correction within the testing family of 48 tests (all q ≥ 0.137). Sensitivity analysis using alternative parameterizations confirmed the ε4+ × global ancestry interactions for pTau_217_ and pTau_181_ with a directionally consistent but non-significant effect in the AFR, EUR, AMR ordering (Figure S9, Table S6). In meta-analysis with the independent cohort, the ε4+ × ILR1 interaction for pTau_181_ was directionally consistent across cohorts and significant in the pooled estimate (pooled β=0.046, 95% CI 0.014 to 0.077) but did not meet our replication criterion (Figure S10, Table S7).

**Figure 3:**
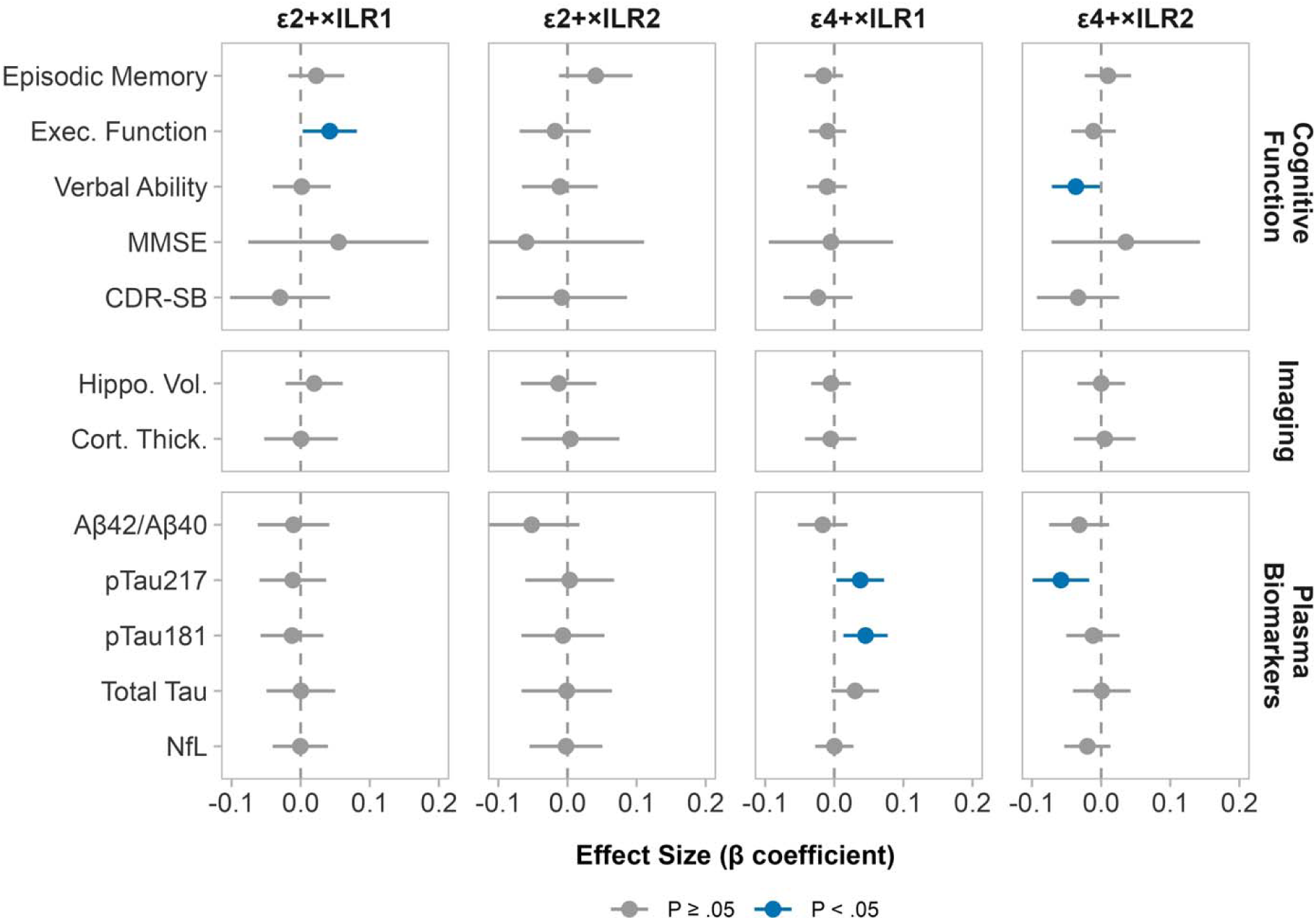
*APOE* × Global Ancestry Proportion Interactions on AD Endophenotypes Using Isometric Log-Ratio Transformation. Interaction effects (β) with 95% CIs showing how global ancestry proportions modulate *APOE* associations with cognition, neuroimaging, and plasma biomarkers. Isometric log-ratio (ILR) coordinates were derived using the (EUR, AFR, AMR) parameterization, where ILR1 represents European versus combined African and Admixed American ancestry (EUR-like vs AFR-like + AMR-like) and ILR2 represents African versus Admixed American ancestry proportion (AFR-like vs AMR-like). Blue points indicate nominal statistically significant interactions (P < .05, uncorrected); gray points indicate non-significant interactions P ≥ .05. No APOE × global ancestry interaction remained significant after FDR correction within this testing family (Table S6). AFR-like, African-like AMR-like, Admixed American-like; EUR-like, European-like ancestry.

### 3.4 Local ancestry modulates *APOE* ε4+ effects on AD endophenotypes

In ancestry-stratified analyses, ε4+ was associated with lower Aβ_42_/Aβ_40_ in EUR/EUR (β=−0.51, 95% CI −0.66 to −0.35) and AFR/AFR (β=−0.51, 95% CI −0.77 to −0.25) individuals but not AMR/AMR. ε4+ effects on pTau_217_ were largest in AMR/AMR (β=0.88, 95% CI 0.47 to 1.27), followed by EUR/EUR (β=0.60, 95% CI 0.46 to 0.74) and AFR/AFR (β=0.20, 95% CI −0.02 to 0.43). ε4+ was associated with higher pTau_181_ in EUR/EUR (β=0.40, 95% CI 0.27 to 0.52) but not AFR/AFR or AMR/AMR.

For cognition, ε4+ was associated with worse CDR-SB, MMSE, episodic memory, and executive function in EUR/EUR individuals, worse CDR-SB and verbal ability in AFR/AFR individuals, and no significant associations in AMR/AMR individuals. For neuroimaging, ε4+ was associated with reduced hippocampal volume in EUR/EUR and reduced cortical thickness in both EUR/EUR and AMR/AMR individuals. AFR/AFR participants showed no significant associations with brain morphometry (Figure 4, Table S8).

**Figure 4:**
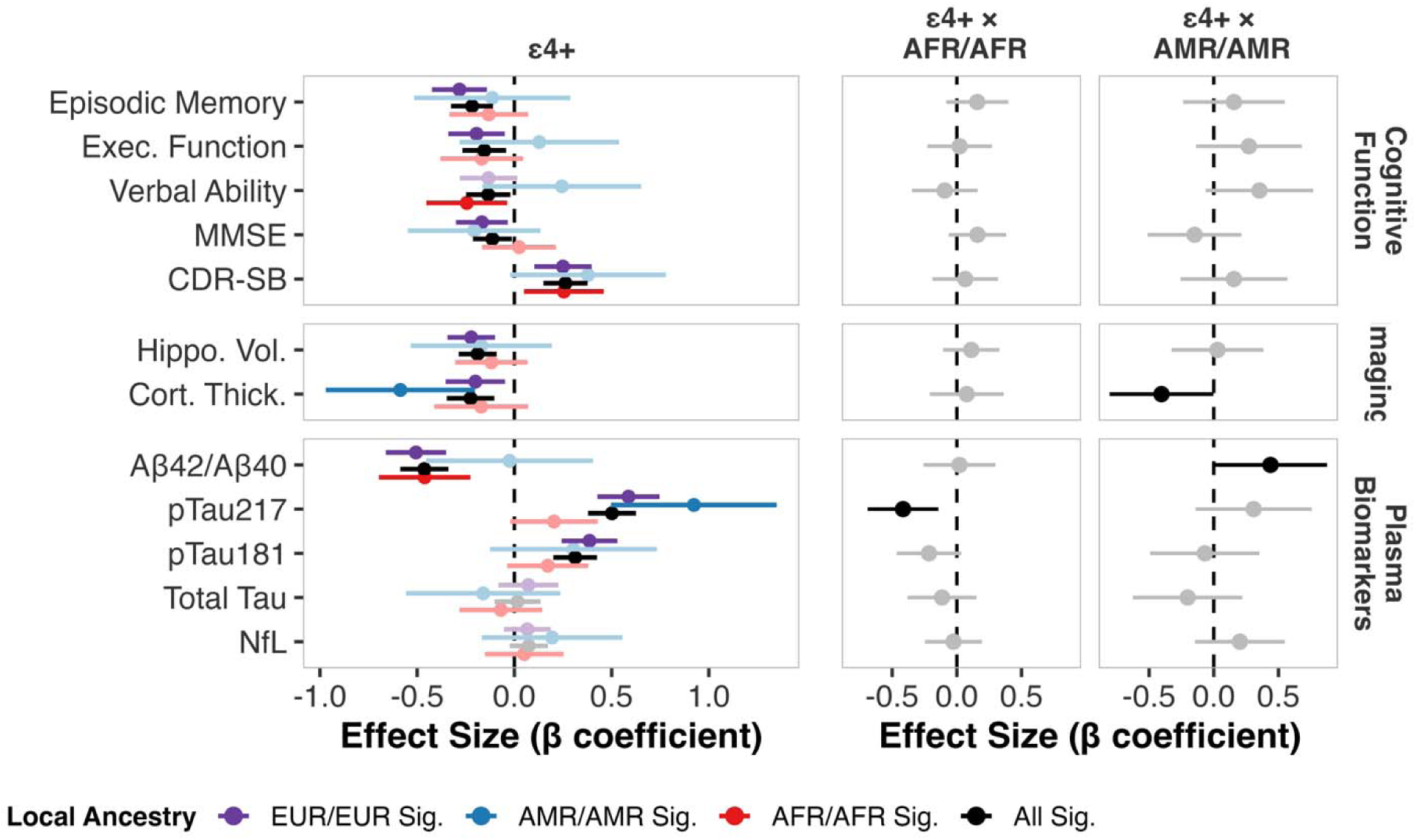
*APOE*\*ε4+ Effects on AD Endophenotypes Stratified by Homozygous Local Ancestry at the *APOE* Locus: Left Panel: Standardized effect sizes (β coefficients, Cohen’s d) with 95% CIs for associations between ε4+ carrier status and AD endophenotypes among participants with homozygous local ancestry at the *APOE* locus. Separate estimates are shown for EUR/EUR (purple), AMR/AMR (blue), and AFR/AFR (red) local ancestry groups. Right Panel: Interaction effects comparing ε4+ associations in AFR/AFR (left panel) and AMR/AMR (right panel) groups relative to EUR/EUR reference group. Filled points indicate nominally statistically significant associations (P < .05); faded points indicate P ≥ .05. No APOE × local ancestry interaction remained significant after FDR correction (Table S9)

Interaction analyses confirmed that ε4+ effects on pTau_217_ were nominally significantly attenuated in individuals with AFR local ancestry at the *APOE* locus (β=−0.42, 95% CI −0.69 to −0.16), and the AMR/AMR cortical thickness reduction was significantly greater than EUR/EUR (β=−0.41, 95% CI −0.82 to −0.004). No significant interactions were observed for cognition (Figure 4, Table S9).

## 4. Discussion

In a community-based cohort of 2,733 participants, we examined how genetic ancestry modulates *APOE* ε4 effects on AD endophenotypes. In the full cohort, ε4+ was associated with lower Aβ_42_/Aβ_40_, higher pTau_181_ and pTau_217_, worse cognitive performance, and reduced brain morphometry. Across all three analytical frameworks, ε4+ effects on pTau_181_ and pTau_217_ were attenuated in participants with African ancestry, while effects on Aβ_42_/Aβ_40_, cognition, and neuroimaging were largely consistent across groups.

Prior studies have documented racial and ethnic differences in AD biomarker levels, including pTau ^5,44,45^, with disparities largely confined to *APOE* ε4 carriers: African Americans show lower tau levels and reduced amyloid burden compared with non-Hispanic Whites, while non-carriers show similar biomarker profiles across racial groups^46,47^. These ε4-driven level differences may have diagnostic implications: although pTau_217_ diagnostic accuracy for amyloid PET positivity is high in both African American (AUC, 0.94) and non-Hispanic White (AUC, 0.92) individuals^48^, uniform thresholds may still misclassify ε4 carriers with African ancestry, where absolute pTau levels are lowest.

However, self-reported race is an imprecise proxy for genetic ancestry, conflating genetic background with social and environmental factors^11^. Indeed, in admixed populations, greater African ancestry is associated with lower neuritic plaque burden and attenuated ε4 effect on neuropathology^49^, consistent with reduced ε4-associated disease risk in populations of African and Amerindian descent^6^. By examining genetic ancestry at continental, global, and local levels, we demonstrate that African ancestry modifies *APOE* ε4-effects on plasma pTau_181_ and pTau_217_.

The ancestry-specific effects on pTau reflect an early stage of AD pathogenesis. Plasma pTau_181_ and pTau_217_ increase after amyloid accumulation, capturing amyloid-associated tau phosphorylation rather than later-stage tau aggregation^50,51^. As T1 markers in the revised AT(N) framework, phosphorylated tau biomarkers (pTau_181_, pTau_217_, pTau_231_) lie downstream of amyloid and upstream of neurodegeneration^52^, positioning them at a stage where *APOE* ε4 primarily influences early amyloid accumulation^28,53^. Our finding of attenuated pTau and pTau_217_ in ε4+ carriers with African-like ancestry is consistent with ancestry-related differences emerging at an early, amyloid-linked stage of the disease cascade.

This interpretation may explain why ancestry-related effects were detectable for pTau_181_ and pTau_217_ but not for plasma Aβ_42_/Aβ_40_, despite amyloid being upstream of pTau in the A/T/N cascade. Plasma Aβ_42_/Aβ_40_ concentrations can be up to tenfold lower in blood than in CSF, largely due to peripheral production from non-cerebral tissues that dilute the brain-derived signal^54^ – making plasma pTau and pTau better suited to detect ancestry-modified *APOE* ε4 effects.

Beyond plasma biomarkers, no ancestry-specific effects were observed in cognitive performance or brain morphometry. Unlike pTau, these outcomes are influenced by multiple non-AD factors, including cerebrovascular disease^55^, educational attainment^56^, cognitive reserve^57^, and measurement variability, which likely reflect greater outcome heterogeneity. Cognitive performance and brain morphometry also reflect later stages of the cascade, where *APOE* ε4 has less direct influence^28,53^, reducing sensitivity to detect ancestry-specific *APOE* effects in a cross-sectional sample. Such differences may be more evident in longitudinal analyses that capture within-person change over time.

Regulatory variation at the *APOE* locus likely represents the primary biological mechanism^13,58,59^. Single-nucleus RNA sequencing shows that ε4 homozygotes with European local ancestry express higher *APOE* levels than those with African local ancestry^58^, suggesting that ancestry-specific regulatory variants influence *APOE* expression. Reduced *APOE* expression in African ancestry ε4 carriers may attenuate amyloid accumulation and consequently downstream tau phosphorylation, yielding lower pTau levels even in the absence of detectable differences in plasma Aβ_42_/Aβ_40_.

Beyond genetic ancestry, other clinical and environmental factors may contribute to ancestry-related *APOE* biomarker differences. Cardiometabolic conditions, which are more prevalent in African American and Hispanic populations^60^, may compound these effects: *APOE*-regulated cholesterol metabolism directly affects amyloid beta processing, and vascular risk factors may interact with *APOE* genotype to influence cerebrovascular pathology^61^. Kidney function, which varies across populations and affects biomarker clearance, may further confound plasma tau and amyloid measurements^43,62^. Psychosocial factors such as stress and social support also differ across populations and have been associated with AD biomarker levels^21^. However, the relative contributions of genetic ancestry, cardiometabolic health, and environmental exposures to ancestry-specific *APOE* effects warrant further investigation.

The study has several limitations. First, the cross-sectional design precludes assessment of how AD endophenotypes change over time. Second, we did not account for educational quality, socioeconomic indicators, vascular risk profiles, or white matter hyperintensities, which could modify *APOE*-biomarker associations and partly explain the observed ancestry-related differences. Third, limited environmental variation within ancestry groups may restrict generalizability to populations with different socioeconomic and environmental exposures. Finally, local ancestry analyses were restricted to concordant homozygous local ancestry (EUR/EUR, AFR/AFR, AMR/AMR), precluding examination of admixed local haplotypes and rare South/East Asian haplotype assignments were reassigned to European/Admixed American ancestry based on shared continental structure.

Despite these limitations, the study has several strengths. First, the comprehensive assessment of multiple AD endophenotypes enabled pathway-specific characterization of ancestry-specific *APOE* ε4+ effects. Second, the use of genetic ancestry measured at group, global, and local levels—rather than self-reported race—provided precise quantification of ancestry-*APOE* relationships validated in an independent cohort. Third, community-based recruitment of underrepresented populations enhances generalizability and addresses longstanding gaps in AD biomarker research.

In conclusion, genetic ancestry moderates the effect of *APOE* ε4 on AD endophenotypes, with African ancestry attenuating ε4+ associations with pTau_181_ and pTau_217_ while Aβ_42_/Aβ_40_ associations are similar. As blood-based biomarkers enter clinical practice, these findings have implications for blood-based diagnostics and motivates evaluation of whether *APOE* genotype and genetic ancestry should be considered in the development and calibration of pTau-based diagnostic algorithms. Current pTau-based diagnostic algorithms do not incorporate ancestry-adjusted thresholds, potentially raising the possibility of differential classification among ε4 carriers with African ancestry who may exhibit attenuated pTau responses despite underlying amyloid pathology

## Acknowledgements

HABS-HD STUDY TEAM HABS-HD MPIs: Sid E. O’Bryant, Kristine Yaffe, Arthur Toga, Robert Rissman, and Leigh Johnson; and the HABS-HD Investigators: Meredith Braskie, Kevin King, James R. Hall, Melissa Petersen, Raymond Palmer, Robert Barber, Austin Reynolds, Yonggang Shi, Fan Zhang, Rajesh Nandy, Roderick McColl, David Mason, Bradley Christian, Nicole Philips, Stephanie Large, Joe Lee, Badri Vardarajan, Monica Rivera Mindt, Amrita Cheema, Lisa Barnes, Mark Mapstone, Annie Cohen, Amy Kind, Ozioma Okonkwo, Raul Vintimilla, Zhengyang Zhou, Michael Donohue, Rema Raman, Matthew Borzage, Michelle Mielke, Beau Ances, Ganesh Babulal, Jorge Llibre-Guerra, Carl Hill, and Rocky Vig.

## Authorship Contributions

SJA has full access to all data in the study and takes responsibility for the integrity and accuracy of the data analysis.

**Concept and design:** Andrews, Boeriu

**Acquisition, analysis, or interpretation of data:** Andrews, Boeriu, Gu

**Drafting of the manuscript:** Andrews, Boeriu

**Critical review of the manuscript for important intellectual content:** all authors

**Statistical analysis:** Fullerton-Howard, Gu, Boeriu

**Obtained funding:** Andrews

**Supervision:** Andrews

## Funding Sources

SJA receives support from the Alzheimer’s Association (ABA 969581) and NIH-NIA (R01AG069453)

AIB receives support from the Alzheimer’s Association (ABA 969581).

EML receives support from NIH-NIA (K00AG080847)

## Conflicts of Interest

The authors report no relevant conflicts of interest.

## Consent Statement

All participants provided written informed consent. The study protocol was approved by the North Texas Regional Institutional Review Board.

## Data Availability Statement

HABS-HD: The data has been made broadly available to the research community. Raw HASB_HD imaging files are stored and processed at LONI (https://ida.loni.usc.edu/). For more information regarding access to HABS-HD.

Analysis code and data processing pipelines are publicly available in four GitHub repositories: apoeDatabase (centralized data processing pipeline for standardizing HABS-HD clinical, genetic, biomarker, and neuroimaging data), apoeAncestry (statistical analyses and manuscript-specific code), adRpheno (AD endophenotype processing including biomarker standardization, cognitive scoring, and APOE genotype classification), and adRutils (R utility functions for data cleaning, transformation, and statistical modeling) (https://github.com/anaboeriu14).

## Additional Information

During the preparation of this manuscript, Claude Sonnet 4.6 was used on the manuscripts to assist in refining the manuscript text for copy editing (clarity, logical flow, and grammar) as well as formatting text to meet style or submission guidelines. The final content, interpretation, and conclusions are solely the responsibility of the authors.

## Notes

### Competing Interest Statement

The authors have declared no competing interest.

### Author Declarations

Ethics committee/IRB of North Texas Regional Institutional Review Board gave ethical approval for this work.

### Summary of Updates

Revised abstracts, methods, and results

