## Supplementary Methods & Figures for "Ancestry-specific effects of *APOE* on Alzheimer Disease Endophenotypes"

**Supplemental Online Content**

Supplementary Methods

Table S1: Distribution of *APOE* ε4 Carrier Status by Homozygous Local Ancestry at the *APOE* Locus

Table S2: Pairwise Comparisons of Baseline Characteristics Across Continental Ancestry Groups

Table S3. *APOE* effects on AD endophenotypes in the entire cohort and stratified by continental ancestry

Table S4: Statistical comparisons of *APOE* effect sizes between ancestry groups (Wald-tests)

Table S5: Meta-Analysis of *APOE* Associations with Plasma Biomarkers Across Continental Ancestry Groups

Table S6: Sensitivity Analysis of *APOE* × Ancestry Composition Interactions Across Alternative Isometric Log-Ratio Transformation Hierarchies

Table S7: Meta-Analysis of APOE × Global Ancestry Proportion Interactions on Plasma Biomarkers

Table S8: Local Ancestry *APOE* Main Effects on AD Endophenotypes

Table S9: Local Ancestry *APOE**ε4+ Interaction Effects on AD Endophenotypes

Figure S1. Cognitive Composite Score Derivation Workflow

Figure S2. Biomarker Harmonization Pipeline for Batch Effect Correction

**Figure S3: Reference Panel Cluster Label Assignments**

Figure S4A: ADMIXTURE Cluster Proportions in (AFR, AMR, EUR) Reference Populations

Figure S4B: ADMIXTURE Cluster Proportions in (EAS, MID, OCE, SAS) Reference Populations

Figure S5: Geographic Distribution of Reference Panel Clusters

Figure S6: ADMIXTURE Cross-Validation Error Across Ancestry Components (K=2 -24)

Figure S7. Concordance between global and local ancestry estimates.

Figure S8A -8C: Meta-Analysis of *APOE* Variant Effects on Plasma Biomarkers by Genetically Inferred Population Groupings

Figure S9: Sensitivity Analysis of *APOE* × Ancestry Composition Interactions Using Alternative Isometric Log-Ratio Hierarchies

Figure S10: Meta-Analysis of *APOE* × Global Ancestry Proportion Interactions on Plasma Biomarkers

**Supplementary Methods**

**Genome-wide genotyping and imputation**

DNA was extracted using the Mag-Bind Blood & Tissue DNA HDQ 96 Kit (Omega Bio-tek) on a Hamilton Microlab STARlet liquid handler and genotyped on the Illumina Global Screening Array (GSA) based on Infinium HTS chemistry. Genotype calls were generated in Genome Studio 2.0 and underwent stringent quality control: variants with call rate <0.95 or Hardy-Weinberg disequilibrium (p < 1 × 10⁻⁶) were removed, as were samples with call rate <0.95, discordant sex based on X chromosome heterozygosity, or excessive/insufficient heterozygosity. Cryptic relatedness was assessed using KING^1^, and one individual from each pair was removed at IBD >0.1875, corresponding to less than halfway between second- and third-degree relatives. Each ancestry group was then imputed separately on the TOPMed Imputation Server using all ethnicities of the TOPMed R3 reference panel, with Eagle for phasing and Minimac3 for imputation^2^. Post-imputation filters removed poorly imputed variants (r² < 0.3), rare variants (MAF < 0.01), and low call-rate variants and samples (<95%) before merging ancestry groups. The final dataset comprised 3,232 samples with approximately 22 million high-quality SNPs per individual.

**Genetically Inferred Population Groups**

Study genotypes were merged with the gnomAD 3.1.2 1000 Genomes and HGDP reference panel, after removing related reference samples, and LD-pruning at r² < 0.2 (200-kb window, 25-kb step)^2^. PCA was performed in PLINK on the reference samples, and HABS-HD individuals were projected onto that space. Distance was computed to the geometric median of each superpopulation (Figure S3).

**Global Ancestry Proportions**

Variants were LD-pruned (100-kb window, r² < 0.1) and ADMIXTURE was run in unsupervised mode across K = 2 – 24 in the reference panel. Cross-validation error declined substantially through approximately K=10–12 and subsequently plateaued; K=12 was selected as a parsimonious representation of population structure because additional components provided minimal improvement in cross-validation error (Figure S6). Participant genotypes were then projected onto these profiles to obtain per-participant ancestry proportions. gnomAD-flagged samples and ancestry-discordant samples were excluded from the analytic dataset, and Middle Eastern reference samples were excluded from the reference panel due to genetic similarity with European and South Asian clusters (Figure S4).

**Local Ancestry Inference at the *APOE* Locus**

G-Nomix^3^ inference was performed using a reference panel of unadmixed 1KG and HGDP individuals with ≥ 95% primary ancestry from one of five superpopulations (AFR-like n =817 , AMR-like n =55, EUR-like n =423, SAS-like n =146, and EAS-like n =718). Since G-Nomix assigns genetic ancestry to each chromosomal segment independently, multiple assignments can occur across the 1Mb *APOE* region within a single haplotype for a given individual. A per-haplotype consensus was then computed: haplotypes with a single consistent ancestry across all segments were assigned that ancestry, while haplotypes with discordant segment-level assignments were labeled as mixed and removed.

**Detailed Statistical Analyses**

*Ancestry Stratified Wald Tests*

The Wald test statistic is calculated as

$$Z=\frac{\beta_{1}- \beta_{2}}{\sqrt{SE_{1}^{2}+SE_{2}^{2}}}$$

where β₁ and β₂ are the regression coefficients from two ancestry groups and SE₁ and SE₂ are their respective standard errors. The test statistic follows a standard normal distribution under the null hypothesis^4,5^ that *APOE* effects are homogeneous across ancestry groups. Pairwise comparisons were conducted between all three groups (AFR-like vs. EUR-like, AFR-like vs. AMR-like, EUR-like vs. AMR-like. Both uncorrected and FDR-corrected p-values are reported in Table S4.

*Compositional Linear Regression with Isometric Log-Ratio Transformation*

Ancestry proportions sum to one, creating a compositional constraint. Including all three proportions as predictors leads to perfect multicollinearity, whereas modeling them independently ignores the inherent trade-off between ancestries — an increase in one proportion results in a decrease in the others.

The isometric log-ratio (ILR) transformation resolves both issues by converting three interdependent ancestry proportions into two uncorrelated, unconstrained coordinates.

$$ILR_{1}=\sqrt{\frac{2}{3}}\times log(\frac{c_{1}}{\sqrt{c_{2}\times c_{3}}})$$

$$ILR_{2}=\sqrt{\frac{1}{2}}\times log(\frac{c_{2}}{c_{3}})$$

Where c1, c2, and c3 represent ancestry proportions in the specified order. The primary analysis used the order of EUR, AFR, and AMR, yielding contrasts of:

- ILR_1_ = EUR vs (AFR + AMR)
- ILR_2_ = AFR vs AMR

Sensitivity analysis tested two alternative orders:

- **(AFR, EUR, AMR):** ILR₁ = AFR vs (EUR + AMR); ILR₂ = EUR vs AMR
- **(AMR, AFR, EUR):** ILR₁ = AMR vs (AFR + EUR); ILR₂ = AFR vs EUR

For the (AMR, AFR, EUR) parameterization, ILR₁ was not interpreted because it conflates AFR and EUR proportions — individuals with identical AMR proportions but opposite AFR-EUR balances receive identical scores despite distinct ancestry profiles. ILR₂ (AFR vs EUR) remained interpretable and was reported, providing a sensitivity check on the AFR-versus-EUR contrast tested in the primary analyses.

*APOE*-by-ILR interactions were tested as:

$$Y= \beta_{0}+ \beta_{1}\left( APOE \right)+ \beta_{2}\left( {ILR}_{1} \right)+ \beta_{3}\left( {ILR}_{2} \right)+ \beta_{4}\left( {APOE \times ILR}_{1} \right)+ \beta_{5}\left( {APOE \times ILR}_{2} \right)+covariates+\epsilon$$

*Local Ancestry Analysis*

Interaction models in the full cohort tested whether APOE*e4 effects differed across homozygous local ancestry groups:

$Y= \beta_{0}+ \beta_{1}\left( APOE \right)+ \beta_{2}\left( \mathrm{LA} \right)+ \beta_{3}\left( APOE\times LA \right)+$covariates + ε

*Missing Data Imputation*

Missing values for body mass index (BMI) and creatinine (for estimated glomerular filtration rate (eGFRcr)) were imputed using random forest imputation implemented in the adRutils R package (<https://github.com/anaboeriu14/adRutils>). Random forest imputation was chosen over single-value imputation to preserve variance structure and capture non-linear relationships among predictors. The imputation model included height, weight, waist circumference, kidney disease status, diabetes, cardiovascular disease, hypertension, dyslipidemia, blood sodium, blood potassium, blood albumin, age, sex, and self- reported ethnicity (available for all participants prior to genome-wide ancestry assignment).

**Supplemental Figures**

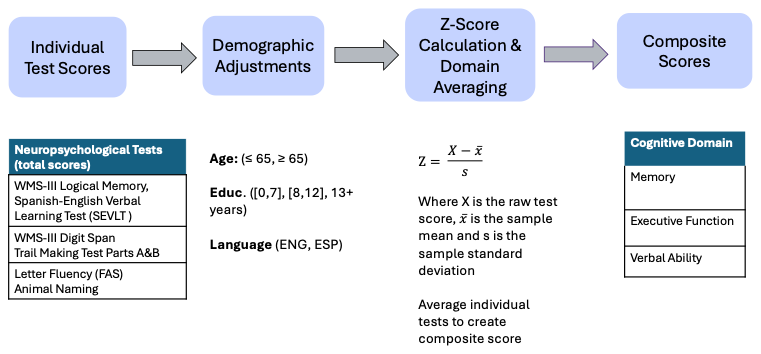

**Figure S1: Cognitive Composite Score Derivation Workflow**

Schematic diagram illustrating the process for creating demographically adjusted cognitive composite scores from individual neuropsychological test scores. Raw test scores are adjusted for age (≤65, ≥66 years), education (0-7, 8-12, ≥13 years), and primary language (English, Spanish). Within each demographic stratum, scores are converted to z-scores using the formula shown, then averaged across tests within each cognitive domain (Memory, Executive Function, Language) to generate domain-specific composite scores.

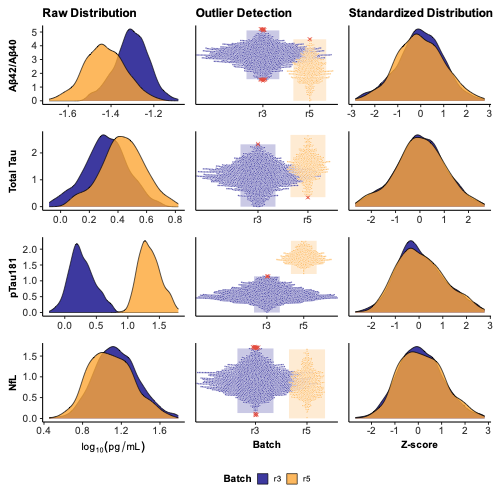

**Figure S2 – Biomarker Harmonization Pipeline for Batch Effect Correction**

Visualization of the plasma biomarker standardization process across three stages: raw distribution (left), outlier detection (middle), and standardized distribution (right) for pTau_181_, total tau, NfL, and Aβ_42_/Aβ_40_ ratio. Raw biomarker concentrations (log10-transformed, pg/mL) show batch-to-batch variability between kit versions (r3 in blue, r5 in orange). Outliers were identified using the interquartile range method (red points indicate values below Q1 - 1.5 × IQR or above Q3 + 1.5 × IQR). Following outlier removal and z-score standardization, distributions from both batches are harmonized with a mean of 0 and a standard deviation of 1, enabling cross-batch comparisons.

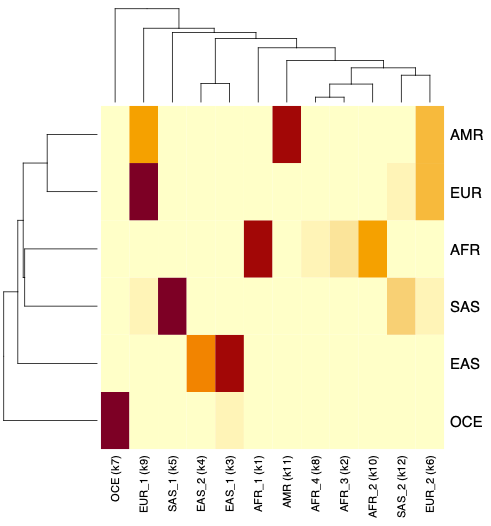

**Figure S3: Reference Panel Cluster Label Assignments**

Heatmap displaying the assignment of K=12 ADMIXTURE clusters to continental super-populations based on mean ancestry proportion. Each row represents a reference super-population (AFR-like, AMR-like, EAS-like, EUR-like, SAS-like) and each column represents an ADMIXTURE cluster. Color intensity reflects the mean proportion of each super-population within a given cluster, with darker colors indicating higher proportions. Cluster labels were assigned to the super-population with the highest mean proportion. European subclusters (EUR_1, EUR_2) correspond to non-Finnish European and Finnish reference samples, respectively. Middle Eastern reference samples were excluded from cluster labeling

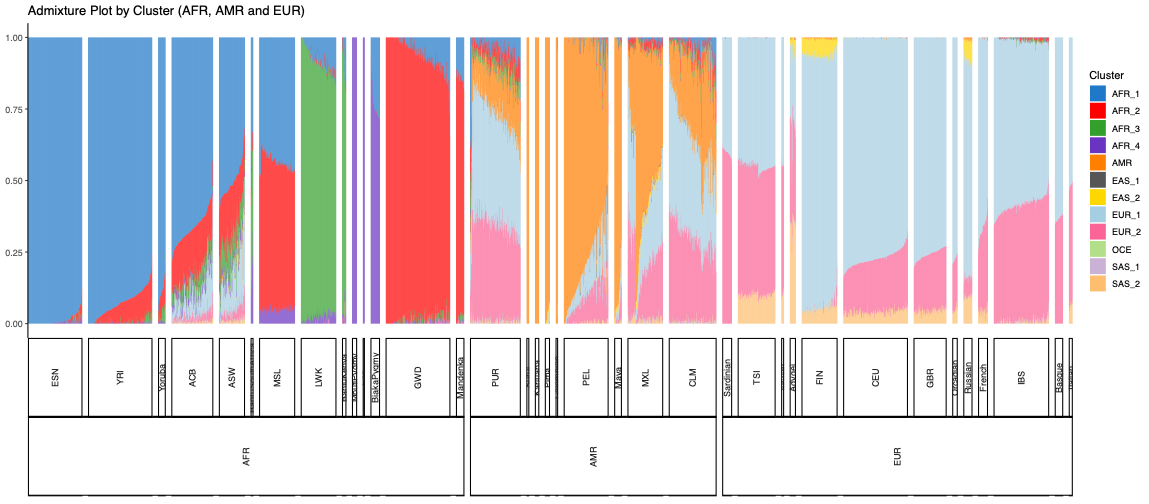

**Figure S4A**: **ADMIXTURE Cluster Proportions in AFR-like, AMR-like, and EUR-like Reference Populations.**

Each vertical bar represents one individual, with colors indicating proportional contributions from 12 ancestry clusters. Populations are grouped by continental superpopulation (AFR-like, AMR-like, EUR-like) along the x-axis. AFR-like, African ancestry; AMR-like, Admixed American ancestry; EUR-like, European ancestry.

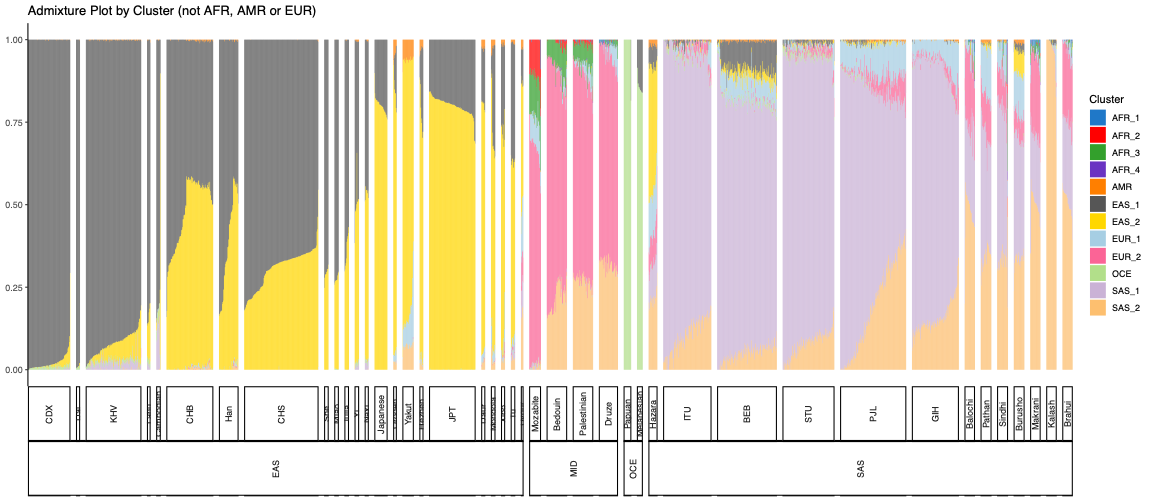

**Figure S4B: ADMIXTURE Cluster Proportions in EAS**-like**, MID**-like**, OCE**-like **and SAS**-like **Reference Populations.**

Each vertical bar represents one individual, with colors indicating proportional contributions from 12 ancestry clusters. Populations are grouped by continental superpopulation (EAS-like, MID-like, OCE-like, SAS-like) along the x-axis. EAS-like, East Asian ancestry; MID-like, Middle Eastern ancestry; OCE-like, Oceanic ancestry; SAS-like, South Asian ancestry.

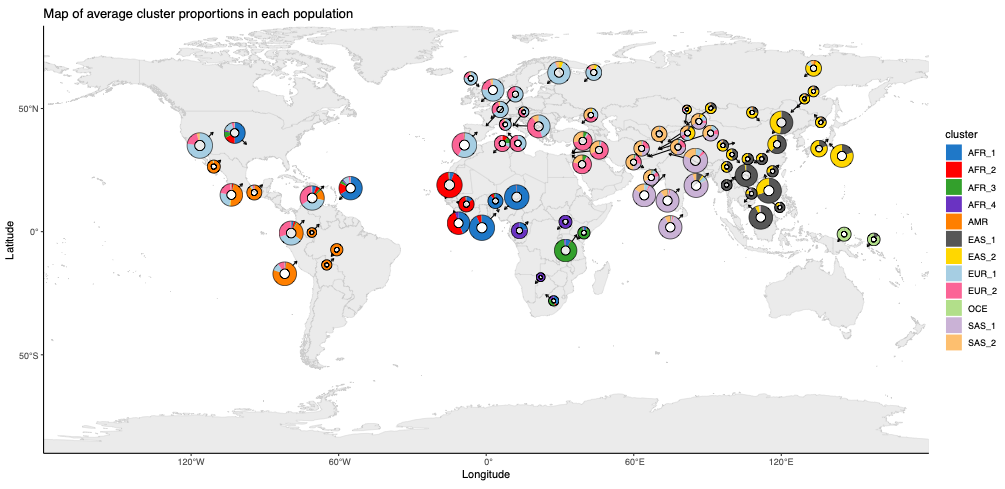

**Figure S5: Geographic Distribution of Reference Panel Clusters**

World map displaying the geographic distribution of average K=12 ADMIXTURE cluster proportions across reference populations from the 1000 Genomes Project and Human Genome Diversity Project (HGDP). Each circle represents a reference population, with pie chart segments colored by cluster assignment. Geographic clustering of ancestry-specific clusters confirms expected continental population structure and supports the validity of super-population label assignments used in continental and global ancestry analyses. AFR-like, African ancestry; AMR-like, Admixed American ancestry; EAS-like, East Asian ancestry; EUR-like, European ancestry; SAS-like, South Asia ancestry.

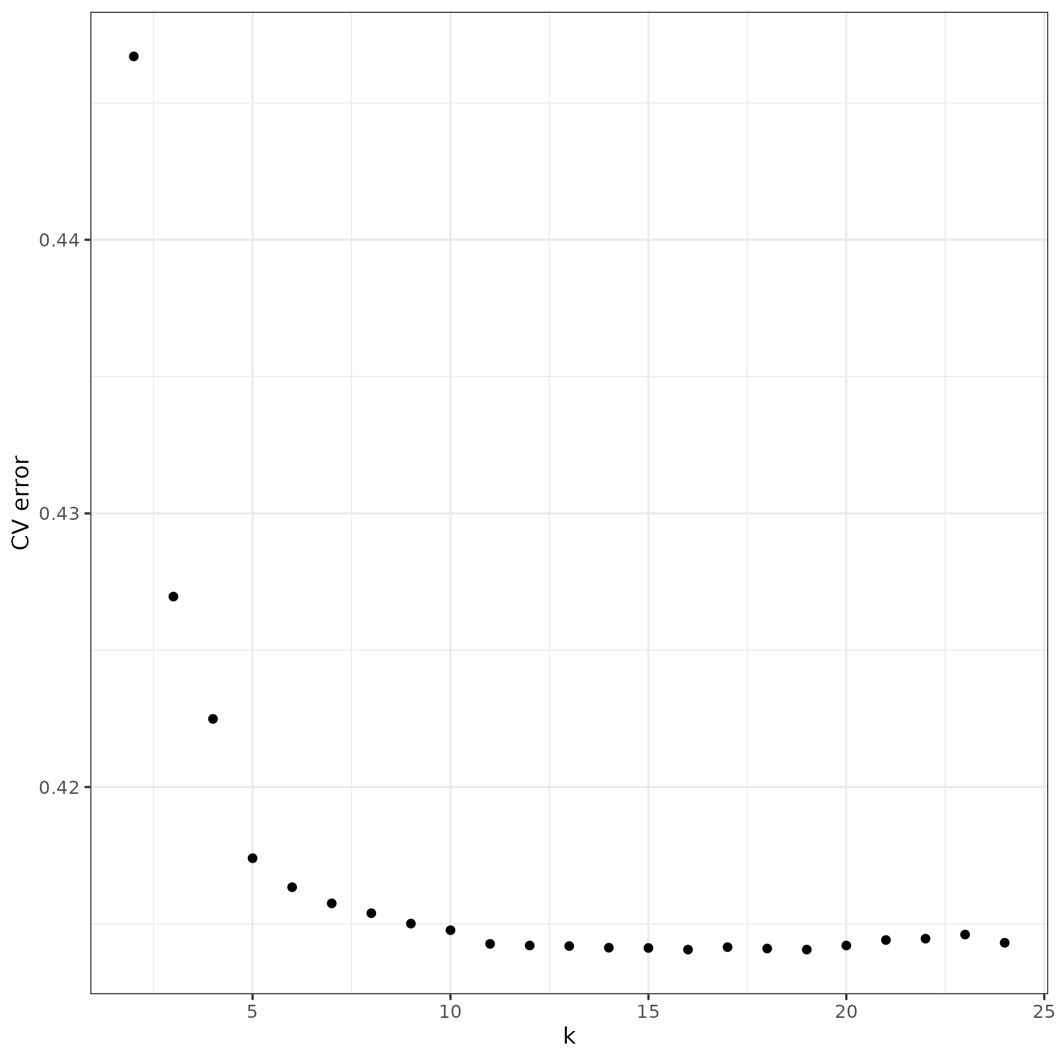

**Figure S6: ADMIXTURE Cross-Validation Error Across Ancestry Components (K=2 -24)**

Cross validation (CV) error from unsupervised ADMIXTURE analysis plotted across a range of ancestry components. Lower CV error indicates better model fit. The curve shows a steep decline from K = 2 to K= 10, followed by a plateau beginning around K=12. Any k beyond 12 represents minimal improvement in the model, indicating minimal improvement in model fit. K = 12 was selected for downstream estimation of global ancestry proportions based on this inflection point. Each point represents the CV error for a single K value.

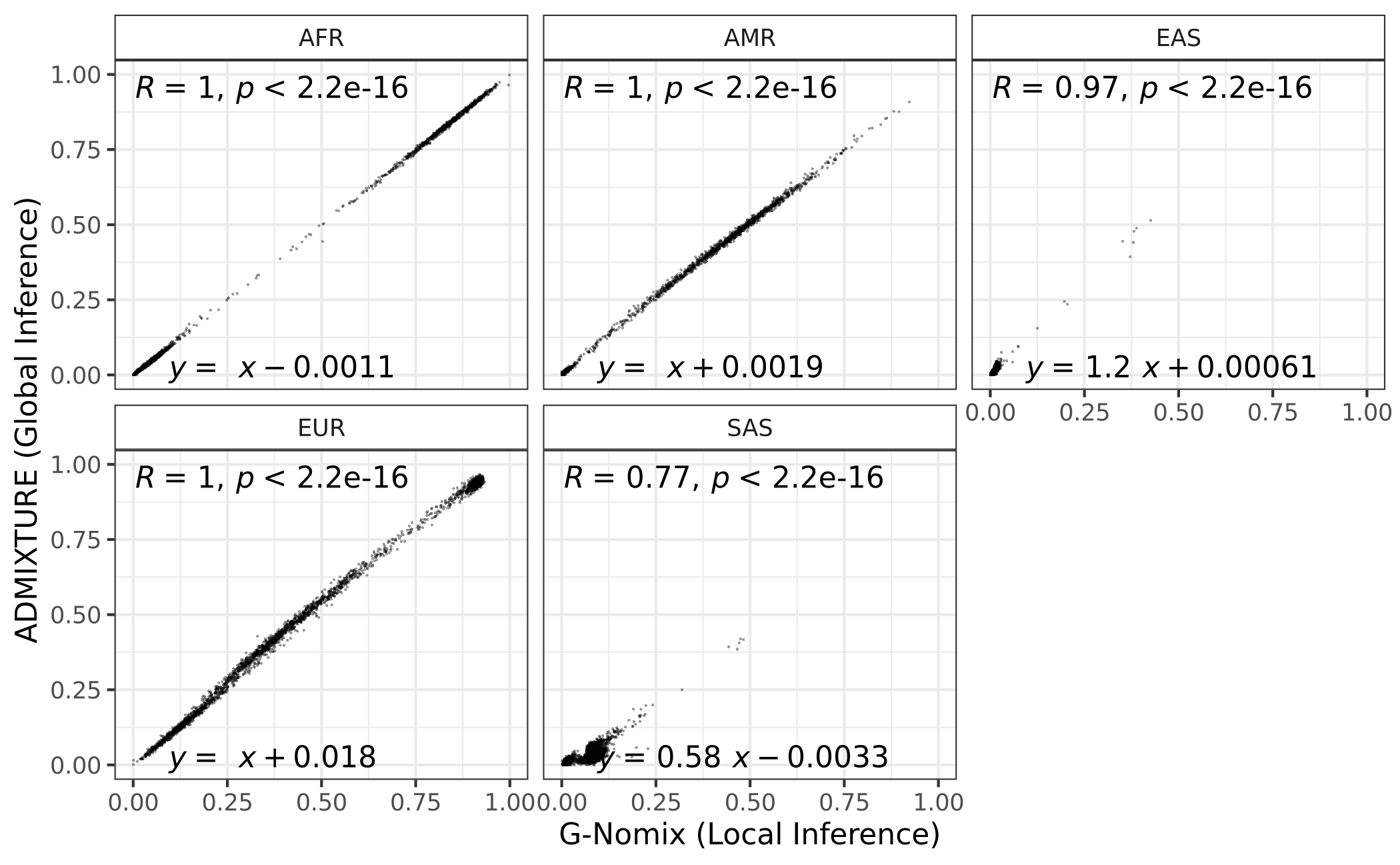

**Figure S7. Concordance between global and local ancestry estimates.** Genome-wide ancestry proportions derived from G-Nomix local ancestry inference were calculated by averaging ancestry assignments across local ancestry segments, weighted by genetic map distance, and compared with global ancestry proportions estimated using ADMIXTURE.

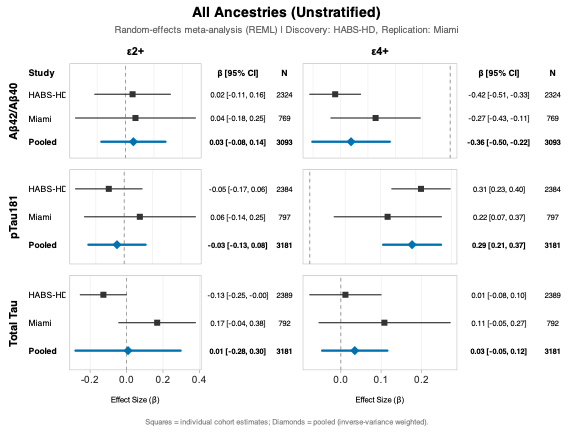

**Figure S8A:** **Meta-Analysis of APOE Effects on Plasma Biomarkers.** Forest plots showing standardized effect sizes (β) with 95% confidence intervals for APOE ε2+ and ε4+ associations with Aβ_42_/Aβ_40_, pTau_181_, and total tau in participants from HABS-HD and Miami cohorts. Squares represent individual cohort estimates weighted by inverse variance; diamonds represent pooled random-effects estimates. Meta-analysis was conducted using REML estimation. ε2+ and ε4+ effects are coefficients from a single model (reference: ε3/ε3); N reflects the full model sample and is shared across both contrasts.

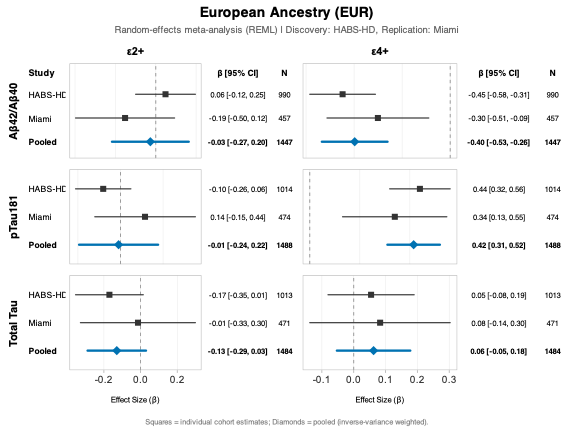

**Figure S8B. Meta-Analysis of APOE Effects on Plasma Biomarkers in European Ancestry (EUR) Participants.** Forest plots showing standardized effect sizes (β) with 95% confidence intervals for APOE ε2+ and ε4+ associations with Aβ_42_/Aβ_40_, pTau_181_, and total tau in EUR-like participants from HABS-HD and Miami cohorts. Squares represent individual cohort estimates weighted by inverse variance; diamonds represent pooled random-effects estimates. Meta-analysis was conducted using REML estimation. ε2+ and ε4+ effects are coefficients from a single model (reference: ε3/ε3); N reflects the full model sample and is shared across both contrasts.

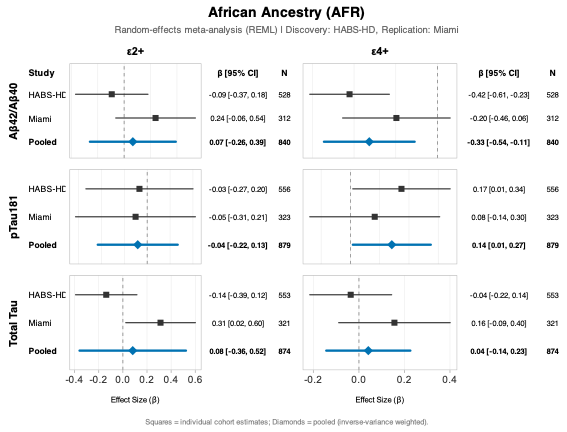

**Figure S8C: Meta-Analysis of APOE Effects on Plasma Biomarkers Across AFR.** Forest plots showing standardized effect sizes (β) with 95% confidence intervals for APOE ε2+ and ε4+ associations with Aβ_42_/Aβ_40_, pTau_181_, and total tau across AFR-like participants from HABS-HD and Miami cohorts. Squares represent individual cohort estimates weighted by inverse variance; diamonds represent pooled random-effects estimates. Meta-analysis was conducted using REML estimation. ε2+ and ε4+ effects are coefficients from a single model (reference: ε3/ε3); N reflects the full model sample and is shared across both contrasts.

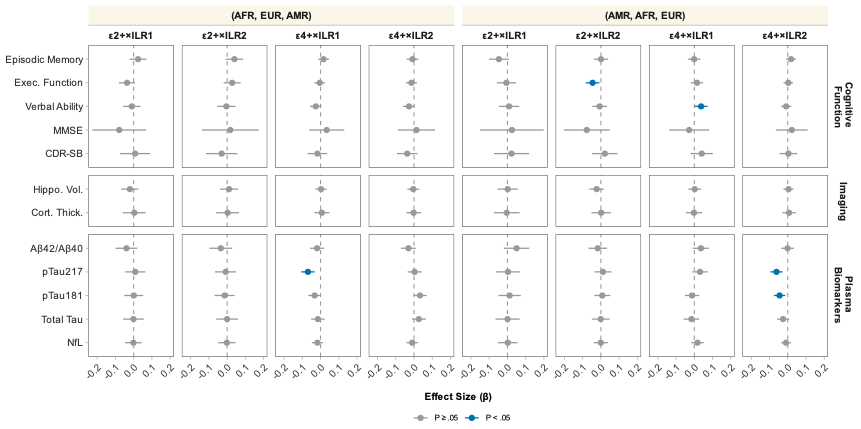

**Figure S9. Sensitivity Analysis of *APOE* × Ancestry Composition Interactions Using Alternative Isometric Log-Ratio Hierarchies**

Forest plots of *APOE* genotype (ε2+, ε4+) × ancestry composition interactions on AD endophenotypes across two isometric log-ratio (ILR) hierarchies. For (AFR-like, EUR-like, AMR-like), ILR1 contrasts AFR-like vs (EUR-like + AMR-like) and ILR2 contrasts EUR-like vs AMR-like; for (AMR-like, AFR-like, EUR-like), ILR1 contrasts AMR-like vs (AFR-like + EUR-like) and ILR2 contrasts AFR-like vs EUR-like. Points represent effect sizes (β) with 95% confidence intervals across cognitive, imaging, and plasma biomarker outcomes. Blue points indicate *P* < .05; gray points indicate *P* ≥ .05. AFR-like, African ancestry; AMR-like, Admixed American ancestry; EUR-like, European ancestry

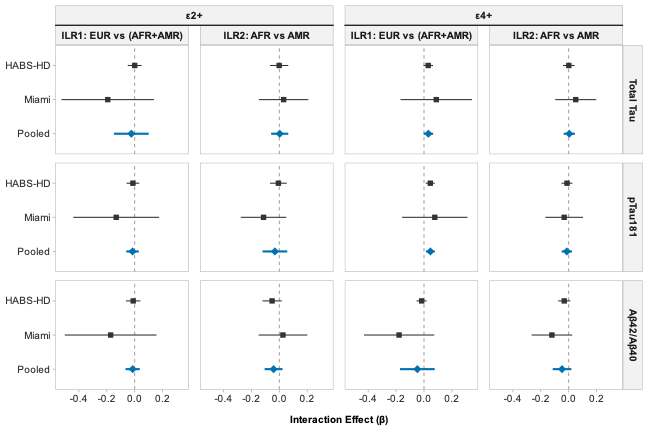

**Figure S10. Random-Effects Meta-Analysis of *APOE* × Global Ancestry Composition Interactions on Plasma Biomarkers Using Isometric Log-Ratio Transformation (EUR, AFR, AMR Parameterization)**

Forest plots of pooled *APOE* genotype (ε2+, ε4+) × ILR ancestry coordinate interaction effects (β, 95% CIs) on plasma biomarkers (Aβ_42_/Aβ_40_, pTau_181_, Total Tau) across HABS-HD and Miami cohorts. ILR1 contrasts EUR vs (AFR+AMR); ILR2 contrasts AFR vs AMR. Squares represent individual cohort estimates; diamonds represent pooled inverse-variance weighted estimates from random-effects meta-analysis (REML). AFR, African-like ancestry; AMR, Admixed American-like ancestry; EUR, European-like ancestry; ILR, isometric log-ratio; REML, restricted maximum likelihood.
